# Methodological challenges and recommendations for identifying childhood immunisations using routine electronic health records in the United Kingdom

**DOI:** 10.1101/2023.02.28.23286573

**Authors:** Anne M Suffel, Jemma L Walker, Colin Campbell, Helena Carreira, Charlotte Warren-Gash, Helen I McDonald

## Abstract

Routinely collected electronic health records (EHR) offer a valuable opportunity to carry out research on immunisation uptake, effectiveness and safety, using large and representative samples of the population. However, using EHR presents challenges for identifying vaccinated and unvaccinated cohorts. Some vaccinations are delivered in different care settings, so may not be fully recorded in primary care EHR. In contrast to other drugs, they do not require electronic prescription in many settings, which may lead to ambiguous coding of vaccination status and timing. Additionally, for childhood vaccination, there may be other challenges of identifying the study population eligible for vaccination due to changes in immunisation schedules over time, different vaccine indications depending on the context (e.g., tetanus vaccination after exposure) and the lack of full dates of birth in many databases of data confidentiality restrictions.

In this paper, we described our approach to tackling methodological issues related to identifying childhood immunisations in the Clinical Practice Research Datalink (CPRD) Aurum, a UK primary care dataset of EHR, as an example, and we introduce a comprehensive algorithm to support high-quality studies of childhood vaccination. We showed that a broad variety of considerations is important to identify vaccines in EHR and offer guidance on decisions to ascertain the vaccination status, such as considering data source and delivery systems (e.g., primary or secondary care), using a wide range of medical codes in combination to identify vaccination events, and using appropriate wash-out periods and quality checks to deal with issues of over-recording and back dating in EHR.

Our algorithm reproduced estimates of vaccination coverage which are comparable to official national estimates in England. This paper aims to improve transparency, quality, comparability and reproducibility of studies on immunisations.

## 1. Introduction

Vaccinations prevent over 3.5-5 million deaths every year and hence, are one of the most successful public health interventions (1). Nevertheless, the global uptake of childhood immunisations has plateaued in the last decade and only 11 countries met a coverage of at least 90% for all recommended vaccines in 2019 (2). The United Kingdom even showed a decline in vaccination coverage over the last ten years below the recommended WHO target of 95% (NHS Digital, 2021; World Health Organization, 2021). In addition to this trend, many vaccination services were interrupted due to the COVID-19 pandemic with up to 25 million children worldwide who missed out on vaccinations and significantly reducing the MMR vaccine uptake in the UK (Firman et al., 2022; McDonald et al., 2020; World Health Organization, 2021).

Population-based observational studies of vaccination are important to maintain and assure immunisation programmes, including understanding and improving vaccine uptake. Furthermore, high-quality studies of real-world vaccine effectiveness and safety are essential to protect health and build public confidence (5). As the determinants of vaccination are complex and very context-specific (6), detailed population-wide data on vaccine uptake can help to design multicomponent interventions that target disadvantaged populations directly (7).

Electronic health records (EHR) are routinely collected from different health care setting which can cover a large parts of the population (8,9). As vaccines are delivered as part of routine health care, EHR have increasingly gaining importance in vaccine research (10–12). EHR can help to explore inequalities in uptake (13), investigate the impact of changes in vaccination schedules (14,15), and assess real-world vaccination effectiveness and safety (16,17). Understanding the context of data collection and curation is a general challenge for high-quality EHR research as these routine datasets were not collected for research purposes but clinical care and administrative processes (18). This impacts how vaccinations are recorded, where the records are saved and which settings they capture.

In this paper, we will discuss different challenges of identifying vaccination events, present recommendations for addressing these and introduce an algorithm to define vaccination status, demonstrated with an example of a cohort of children born between 2006 and 2014 in England from Clinical Practice Research Datalink (CRPD) Aurum.

## 2. The example cohort

Primary care data sets such as CPRD Aurum provide rich, longitudinal patient-level information from a primary care setting on symptoms and diagnoses, clinical tests and results, immunisations, prescriptions and referrals to other services (9). CPRD Aurum contains anonymised data originally collected from primary care practices in England and Northern Ireland using the EMIS Web® electronic patient record system software to manage patient care. In 2022, CPRD Aurum contained data from around 41 million patients and was broadly representative in geographical spread, age, sex and ethnicity (9,19). Data on clinical diagnoses, symptoms, clinical tests and referrals are collected in an observation table using SNOMED CT, Read Version 2 and local EMIS Web © codes, whereas data on drug prescriptions and devices are collected in a separate drug issue table coded using Dictionary of Medicines and Devices (DM+D) (9). Coded records for clinical diagnoses in CPRD usually show a high validity in CPRD GOLD but less information is available for CPRD Aurum (20).

The data from practices can be usually linked to secondary care data sets, death registries, and either patients’ or local practices’ area-level measures of relative deprivation (9,21).

To illustrate the different steps and challenges of identifying childhood immunisations using EHR, we described the vaccination status for routine childhood vaccination schedule up to the age of five for an exemplar cohort of all children in CRPD Aurum who were born between 2006 and 2014. Each child was followed up until they either changed GP practice, died or turned five years old. To preserve confidentiality, CPRD provides only month and year of birth for children rather than an exact date of birth. Therefore, to better estimate the timing of each vaccination, their date of birth was set to the 15^th^ of their month of birth. After conducting quality checks which entailed using an acceptability flag provided by CPRD, we removed children who had a record of vaccination within two weeks around their estimated date of birth as we interpreted this as indicator of unreliable vaccination recording. The final study population included 1,743,030 individuals.

A summary table of the demographics of the included children can be found in the supplementary material (see table S.1)

## 3. Selecting a data source and study definition of vaccination

### 3.1 Selecting a data source to reflect vaccine delivery

In the UK, routine childhood vaccinations up to the age of five are largely delivered and recorded in primary care (Tiley et al., 2020). However, vaccines can also be delivered in a variety of different settings: for children, this includes schools for children aged 4 years and over, secondary care for high risk groups who are recommended additional vaccines, and pharmacies (see Table 1). Other vaccines, such as travel-related vaccines or the chickenpox vaccine, are mainly given privately in pharmacies or private clinics. Maternal vaccinations in pregnancy can be delivered in primary care or in hospital antenatal clinics (22).

**Table 1.** Overview of different settings in the UK where vaccines are usually administered to children.

| Vaccine | Age group | Typical setting |
| --- | --- | --- |
| 6-in-1 vaccine | Children under age 1 | Primary care |
| Rotavirus vaccine | Children under age 1 | Primary care |
| Hib/MenC | Children under 5 | Primary care |
| MMR | Children under 5 | Primary care |
| 4-in-1 preschool booster | Children under 5 | Primary care |
| MenACWY | Adolescents (13-15 years) | School |
| HPV | Adolescents (12-13 years) | School |
| 3-in-1 teenage booster | Adolescents (13-15 years) | School |
| Influenza | Children | School |
|  | Clinical Risk Groups | Primary Care |
| BCG* | Clinical Risk Groups | Hospital<br>Primary Care |
| Hepatitis B | Clinical Risk Groups | Hospital |
| Chickenpox | Clinical Risk Groups | Secondary Care |
| COVID-19** |  | Primary Care,<br>Pop-up vaccination centres,<br>Pharmacy, Schools |

6-in-1: includes vaccines against diphtheria, hepatitis B, Haemophilus influenza type b, polio, tetanus, and pertussis. Hib: Haemophilus influenza type b. MenC and MenACWY: meningitis. 4-in-1 preschool booster: diphtheria, tetanus, pertussis, polio. HPV: Human papillomavirus. 3-in-1 teenage booster: tetanus, diphtheria, polio. BCG: tuberculosis vaccine. Clinical risk groups: depending on the vaccine, groups of people who are at higher risk of contracting the disease or who are clinically vulnerable due to other health conditions. \*The BCG vaccination has been subject to recent changes, hence it will require more research into the changed delivery systems and quality of available data. \*\*Data on the COVID-19 vaccine is collected in a national register through different point of care apps and automatically feed into the GP records.

As there are a variety of different settings where childhood immunisations can be administered, this implies a careful choice of the data source for a vaccine study. In theory, UK primary care data should include a record of all vaccinations, wherever received. However, the completeness of recording in primary care depends on where the data was collected and its transfer from other vaccinating settings, which are variable in schools (23) and antenatal clinics (22), resulting in incomplete or delayed immunisation records. Vaccinations given in other settings, such as pharmacies, might not be well recorded at all. Depending on the setting where the data was collected, there might be some time lag between vaccine administration and data transfer into the GP record. This should be considered in sensitivity analyses when applying minimum ages to vaccine receipt (see section 5).

For example, for the influenza vaccine, there are several delivery settings. At school age, usually a live attenuated influenza vaccine (LAIV) with nasal application is administered to children (24). However, children who are at increased risk of flu due to underlying health conditions will receive their flu vaccine in a GP practice. Similar applies to children who cannot have the LAIV because of allergies, use of porcine gelatine stabilisers, immunosuppression or severe asthma (25). Ideally, data from schools and GPs should be linked representing all groups of children. If this is not possible, researchers have to be aware of potential misclassification and bias in their study.

#### Recommendations

- *Investigate in what settings the vaccine of interest is usually administered*.
- *Check how well these settings are covered by available datasets*.
- *Use data linkages where possible to cover several settings of vaccine administration*.
- *Consider and discuss misclassification and bias which may arise from the datasets used*.

### 3.2 Ensuring the definition is robust to changes in vaccine schedules over time

The vaccine schedules have been changed and adapted many times over the recent years and will be subject to ongoing changes (UK Health Security Agency, 2013). This especially applies to childhood immunisations. If the researcher is interested in a specific antigen, it is consequently important to choose a study period which did not include any change of schedule for the vaccine of interest or to ensure that the study definition of vaccination is appropriately time-updated to coincide with changes to the vaccine schedule.

If this is not possible or if the general focus is on the childhood immunisation schedule as a whole, we recommend focussing on different antigens which have been continuously given at the same age. For our example cohort, this meant focusing on antigens like tetanus, diphtheria, pertussis, pneumococcus, measles mumps, and rubella as those have been consistently recommended at the same ages between 2006 and 2019.

However, this approach leads to another question which is whether a single antigen can be interpreted as representative for all the vaccines given at the same age. This will be further discussed in section 6.

#### Recommendations

- *Investigate changes in the routine immunization schedule for the vaccine of interest*.
- *Limit study period to a time with no changes in the schedule or stratify your analysis by the time of changes in the schedule*.
- *If interested in the overall immunisation schedule, focus on antigens which have been consistently given throughout the study period as part of a vaccine combination (e*.*g*., *all vaccines with pertussis antigen) and use them to represent different appointment within the schedule*.

## 4. Categorising records of vaccination events within a primary care data source

### 4.1 Prescription records do not fully capture vaccine delivery

In addition to the different settings where a vaccine can be delivered, different methods of vaccine administration can impact the way of how a vaccine recording in a routine health care setting.

If a vaccine is administered in the same practice where the patient is registered, this means no electronic prescription of the vaccine is necessary for dispensation using Patient Group Directions (PGD) and Patient Specific Directions (PSD) (26,27). Hence, using electronic prescription data only would underestimate the number of vaccine doses given in this practice.

We explored this issue in our example cohort. For this purpose, we classified vaccine-related codes in CPRD Aurum into four different categories, such as administered vaccine code, neutral vaccine codes, vaccine product codes and vaccines refusal codes. More detailed information on which codes were used and how they were classified can be found in the supplementary material. Table 2 shows that less than 0.2% of all the extracted codes for different vaccines were codes from the prescription table. Consequently, we recommend combining prescription records with other coded vaccination records in the health data for studies in the UK. Section 4.2 will go into more detail about the code list generation.

**Table 2.**
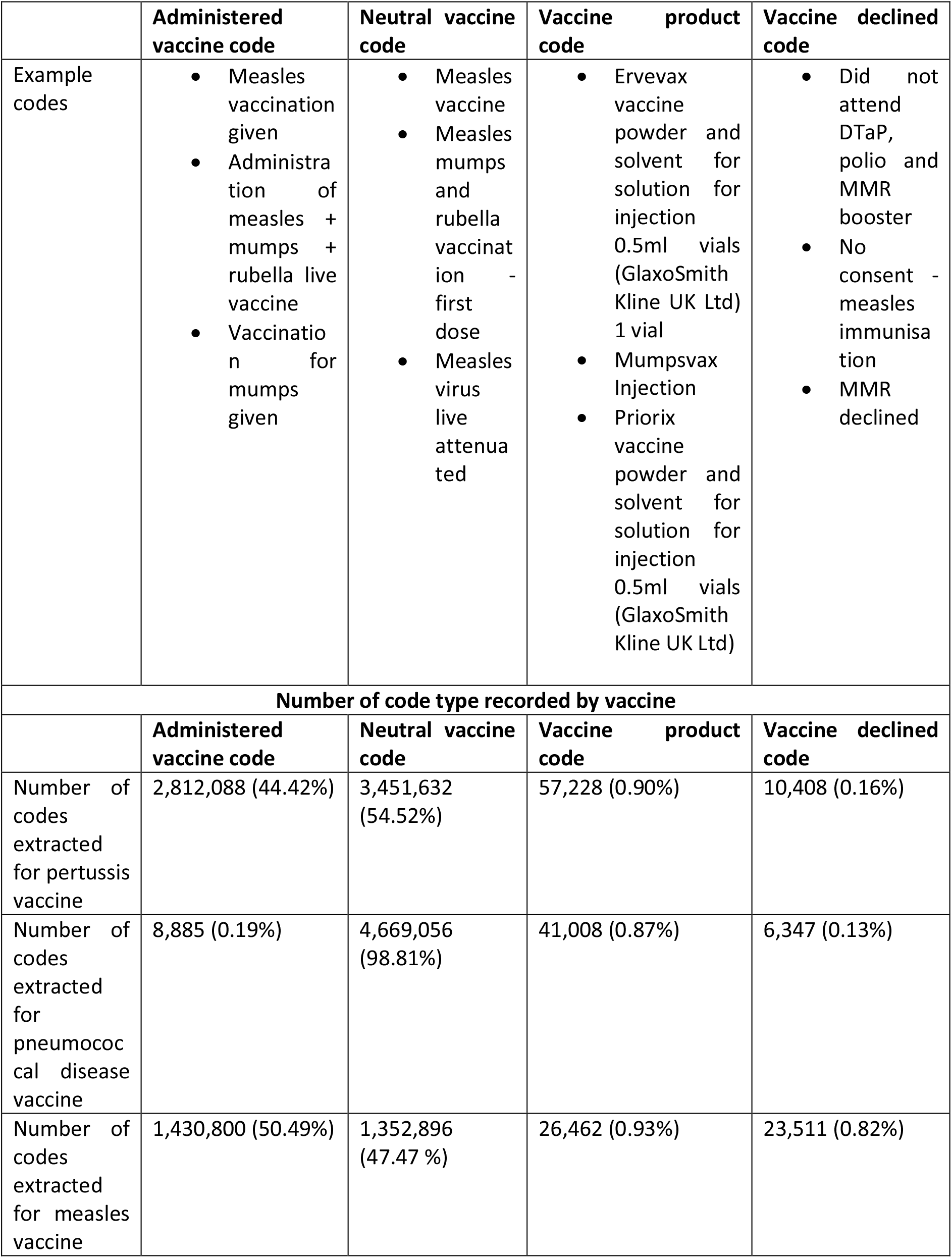
Examples of different codes for the MMR vaccine falling under different categories and the proportion of codes falling under each category by vaccine type.

#### Recommendations

- *Identify all the different ways how vaccination might be encoded in your dataset (e*.*g*., *prescriptions, procedures or events, etc*.)
- *In a UK context, do not use prescription codes only but combine them with event-related codes*
- *Code lists can be found in the appendix*.

### 4.2 Capturing ambiguous or conflicting records of vaccination events

Medical codes related to vaccines can be ambiguous as to whether a vaccine was given or be linked to a declined vaccination, adverse events or just the recall process for parents. They can be very specific (“MMR preschool booster”) or very general (“Childhood Immunisation”). Hence, there are two main considerations to be made: what categories of vaccine codes should be used to define vaccination events and how to deal with combinations of different codes which might be contradictory.

#### 4.2.1 Categories of vaccination codes

Firstly, we investigated the impact of different categories of vaccine codes on defining vaccination events. After exploring the commonly used SNOMED codes in relation to the MMR vaccine, we decided to use the following four categories: codes for an actively given vaccine (indicated by words used such as ‘vaccine given’ or ‘vaccine administered’), neutral vaccination codes (e.g., ‘MMR vaccine’), product codes (from prescription records) indicating a specific pharmaceutical product used, and codes for declining a vaccine. We counted codes just naming the type of vaccine (e.g., ‘measles vaccine’) as neutral as they can be used together with other codes indicating a discussion or invitation to the vaccine. Table 2 shows an example for each of these categories for the MMR vaccine.

We chose not to include any codes about invitations to immunisations or recall as they add no information on whether a vaccine was given. It can be debated whether codes for adverse events after vaccination should be included as they clearly indicate that a vaccine was given at some point. However, we decided against including them into our algorithm as they these codes were rarely used (e.g., only 103 adverse events in 13 years of administering the MMR vaccine) and they added little information on the vaccine timing for the study.

In our cohort, the profile of codes differed by antigen, for example with codes which were ‘neutral’ as to whether the vaccine was given comprising 98% of records of pneumococcal antigen, and approximately half the records for pertussis and measles antigen (Table 2).

A more detailed description on the code lists used and how the codes were classified, can be found in the supplementary material.

#### Recommendations

*A combination of different types of codes capturing vaccination is essential in EHR*

*Focusing on products or codes of administration only will substantially underestimate the number of vaccination events, researchers should also use codes which are neutral as to whether the vaccine was given*

#### 4.2.2 Considering non-specific codes for vaccination events

After identifying different vaccine-related codes, the use of general vaccine codes, such as “childhood immunization” or “vaccine”, needs further consideration. Firstly, this will depend on the overall question of the study as only specific vaccine codes can guarantee that the vaccine of interest was indeed received. Nevertheless, using only specific codes might underestimate the total number of vaccinations when low specificity is required (e.g., in studies using vaccination as proxy for health-seeking behaviour).

To explore this issue, we compared the results for extracting vaccine related data using specific code lists for the DTP vaccine only or using specific codes together with a list of very general vaccine codes as described above. The results can be found in the supplement (Table S.2). Using more general terms detects more different vaccine events falling under different categories with the exception of vaccine products (see Table S.2). However, after applying our vaccine algorithm, the overall number of vaccine doses per person was nearly the same with the exception of a higher number of declined vaccinations for people who had no vaccine record before (see table S.3).

#### Recommendations

- *Due to the low specificity of the general codes, we would not generally recommend using general vaccine codes as they do not add the number of identified vaccination events*.
- *Nevertheless, the general vaccine codes can be helpful for studies specifically looking at declined vaccinations as vaccine refusal seems to be more likely to be coded with a general code than with a vaccine specific code*.

#### 4.2.3 Dealing with conflicting vaccination codes

Records which are neutral as to whether a vaccine was administered should be interpreted differently if they are recorded together with a prescription or a declined vaccination record. To determine if a vaccine had been delivered using the combined records on any given day the following algorithm was applied: for each patient the number of administered, neutral, product and declined codes were summarised by date of the observation. Different combinations of codes led to interpretation of a child being vaccinated, a declined vaccination or conflicting codes. A neutral code might combine with a prescription or vaccine administered code to confirm vaccine delivery that day, or combine with a vaccine declined code to be interpreted as a vaccine not given that day (Table S.4).

Overall, there were very few conflicts observed of a vaccine declined code together with either a prescription or a vaccine administered code (e.g., 327 for the MMR vaccine (<0.01%)). These events were excluded from the analysis. We did not find any instances of three different code combinations on the same day.

This process largely confirmed vaccine delivery dates, but also identified dates of vaccine refusal and a small number of dates with conflicting records for each vaccine (see Table S.5).

#### Recommendations

*Assessing different types of vaccination codes recorded on the same day is important in order interpret the use of neutral vaccination codes correctly and to correctly identify vaccine refusals*.

## 5 Determining the number and timing of a vaccine dose

### 5.1 Summary of the vaccination algorithm

Most vaccines which are part of the NHS childhood immunisation schedule consist of several doses which should be given within a certain time period and usually require a minimum gap between doses to ensure maximum efficacy (28). In order to evaluate the success of vaccination programme it is important to determine how many doses have been administered and whether they have been administered within the correct time interval. In real-world data this is challenging as vaccine dose number is not typically recorded. Furthermore, determining the correct timing of a vaccine with respect to the child’s age is important as some vaccines are not effective when given too early (29) or lead to a higher risk of transmission when delayed (30) or adverse effects from the vaccine (31).

The first difficulty is that it might not be possible to detect the precise age at vaccination in anonymised datasets as the exact day of birth may be not provided, for example being suppressed in CPRD Aurum to protect patient confidentiality (9). We dealt with this issue by setting every child’s day of birth to the 15^th^ of the respective month and then allowing a range of 16 days uncertainty for every interval.

The second difficulty is that after identifying potential vaccination events as described in section 4., it is essential to differentiate separate vaccination events from multiple records of the same event. Particularly by using vaccination codes which we classified as ‘neutral’, there is still a possibility that one of the codes referred only to the discussion of the vaccination or any other more administrative issue or side effects related to the vaccine. These records may result in apparent second doses - for UK childhood vaccinations, the minimum intervals between vaccinations may be as little four weeks (28,32).

For dealing with challenges of multiple reporting of the same event in EHR and short time intervals between vaccines, we developed an algorithm which applied a combination of minimum ages for each dose, and a minimum time interval between different vaccination doses, to define the timing of vaccine doses, illustrated in Figure 2 and described in detail in the supplementary material (see S.6).

As an example for exploring potential minimum age gaps between doses, figure 2 shows the number of vaccine event which would be dropped by applying different minimum age gaps for individuals between doses 1 and 2, and doses 2 and 3 for the pneumococcal vaccine. It reflects the recommended four week gap between two doses and shows that some doses might be caught up one or two weeks later than recommended.

Figure S.8 describes the probability density of recording a vaccination event by different ages on a population level. It shows that especially early doses have be to differentiated carefully as there might be overlap of vaccine doses if definitions are based on age alone. However, all different doses show distinct peak ages of uptake.

Figure 3 shows how many vaccination records were removed at every stage of the algorithm.

Table S.7 in the supplement gives a summary of recommended minimum age and age gaps for different vaccines. After applying this algorithm, there were still some individuals who had more than the four recommended pertussis vaccine doses recorded. 3,654 children had a total of five vaccine doses recorded, 169 children six doses and 16 children had a total of seven doses recorded. All fifth doses and higher were dropped from the data set. A potential explanation for the recording of more doses could be double recording of the same dose, maybe additional doses which were given after exposure to one of the antigens in the combined vaccine product or a potential misclassification of the event itself, i.e., only discussion regarding a vaccination instead of actually administering the vaccine.

#### Recommendations

- *Apply both minimum age thresholds and minimum time intervals between doses to determine timing of vaccine doses*.
- *Consider the importance of timeliness for a vaccine dose when applying these thresholds*.

### 5.2 Results from applying the algorithm to the example cohort

After conducting the vaccine algorithm, we calculated the vaccine coverage at different ages and compared them to national NHS Digital estimates (33) (see Table 3). Our algorithm led to a very similar or even marginally higher vaccine uptake in the study population in comparison to national estimates with exception of earlier years of the pneumococcal vaccine.

**Table 3.** Overall vaccine coverage by calendar year from the vaccination algorithm in comparison to the NHS digital estimates of the same year in England.

| Year | DTP vaccine | National estimates DTP* | Pneumococcal vaccine* | National estimates Pneumococcal* | MMR vaccine coverage | National estimates for MMR* |
| --- | --- | --- | --- | --- | --- | --- |
| 2009-10 | - | 84.8 | 84.25 | 87.6 | - | 82.7 |
| 2010-11 | - | 85.9 | 84.62 | 89.3 | - | 84.2 |
| 2011-12 | 87.03 | 87.4 | 84.17 | 91.5 | 90.04 | 86.0 |
| 2012-13 | 87.99 | 88.9 | 87.05 | 92.5 | 90.78 | 87.7 |
| 2013-14 | 89.00 | 88.7 | 88.80 | 92.4 | 91.56 | 88.3 |
| 2014-15 | 89.42 | 88.5 | 90.29 | 92.2 | 91.23 | 88.6 |
| 2015-16 | 89.46 | 86.3 | 91.42 | 91.5 | 90.50 | 88.2 |
| 2016-17 | 89.21 | 86.2 | 91.53 | 91.5 | 89.91 | 87.6 |
| 2017-18 | 88.91 | 85.6 | - | 91.0 | 89.37 | 87.2 |
| 2018-19 | 88.82 | 84.8 | - | 90.2 | 89.20 | 86.4 |

DTP: diphtheria, tetanus, pertussis vaccine. MMR: measles, mumps, rubella vaccine. \*Coverage for the pneumococcal vaccine was measured for three doses at the age of 2. Coverage for DTP and MMR refer to the completion of the preschool booster (4<sup>th</sup> and 2<sup>nd</sup> dose respectively) at the age of 5.

## 6. How to deal with different antigens recommended at the same vaccine appointment

Vaccines in the routine childhood immunisation schedule are usually given as a combination of different antigens in order to reduce the number of injections for children (28). Depending on the question of interest, researchers might be interested in more than one antigen. Different antigens can be either given together as one combined injection, e.g., measles and mumps vaccine, or they can be given at the same appointment but as separate injections, e.g., measles and pneumococcal vaccine at the age of one year.

We explored for our study cohort whether antigens should be considered separately or if one antigen can represent a vaccination appointment. Table 4 shows the number of events detected using only a single antigen or several antigens which are usually given together as one vaccine.

**Table 4.** Number of vaccination events and patients with vaccination detected by different types of vaccine code list.

|  | Number of vaccine-related events in data set | Number of patients with any vaccine related record |
| --- | --- | --- |
| Code list containing all codes for measles, mumps and rubella antigen containing vaccines | 3,955,205 (100%) | 1,531,523 (100%) |
| Code list containing codes for measles antigen containing vaccine only | 2,894,040 (-26.82%) | 1,531,396 (-0.00%) |
| Code list containing all codes for diphtheria, tetanus and pertussis antigen containing vaccines | 9,058,339 (100%) | 1,702,697 (100%) |
| Code list containing codes for pertussis antigen containing vaccine only | 6,389,450 (-29.46%) | 1,700,423 (-0.00%) |

Although the number of events recorded notably decreases around 27-29% from using all antigens versus only a single one, the overall number of patients with any vaccine record stays roughly the same. To explore whether the reduction of vaccine-related health records using only one vaccine antigen instead of all vaccine antigens leads to a change of estimated vaccination rates after applying the vaccination algorithm above, we calculated age of vaccine receipt and vaccine coverage at different age for the example of the MMR vaccine using these two different code lists approaches. Table 5 illustrates how the results of overall vaccine uptake and timing of vaccine receipt are impacted by using either all antigens or only a single antigen.

**Table 5.** Vaccine uptake and coverage for the MMR vaccine described using two different types of code lists to identify MMR vaccine-related health records.

| Vaccine type | Dose | Mean age of receipt in days | Median age of receipt in days | IQR in days | Coverage at the age of two (in %) | Coverage at the age of five (in %) |
| --- | --- | --- | --- | --- | --- | --- |
| MMR (measles antigen based) | 1 | 455.50 | 409 | 387-446 | 93.86 | 97.55 |
|  | 2 | 1273.00 | 1,269 | 1236-1321 | 2.25 | 90.23 |
| MMR (all antigen based) | 1 | 455.40 | 409 | 387-446 | 93.87 | 97.55 |
|  | 2 | 1272.00 | 1,269 | 1236-1320 | 2.34 | 90.28 |

MMR: measles, mumps, rubella vaccine. IQR: interquartile range.

There are minimal differences in coverage and timing of vaccine receipt for each MMR dose when code lists for either the measles antigen or for all antigens are used. The single antigen estimates are still slightly higher than the NHS Digital estimates for vaccination coverage by calendar year (see Table 3).

### Recommendation

- *A code list for a single antigen is usually sufficient to estimate uptake and coverage for vaccine products combining several antigens*.

## 7. Considerations for vaccinations in special situations

Another challenge for correctly calculating the vaccine coverage is vaccines which are part of the routine immunisation schedule but are sometimes given differently under special circumstances. For example, the DTP vaccine may be given after injuries or other relevant exposure for tetanus infection (34), and the first MMR vaccine is given earlier to high-risk individuals and areas with risk of outbreaks in the UK (29).

For the DTP vaccine, this means that there is a risk of mistaking a post-exposure vaccine for a later vaccine dose. We addressed this issue by applying a minimum age and a minimum age gap for the later DTP dose as a post-exposure vaccine cannot replace the preschool booster (34). If a study focus is post-exposure vaccinations, a different approach has to be chosen.

For the MMR vaccine, more specific considerations have to be made: if a vaccine was given under the age 1, this vaccine cannot replace the first routinely scheduled vaccination (29) and hence, we removed all vaccines given under the age of one in our vaccine algorithm. If a study is looking at vaccination in outbreaks settings and interested in the additional vaccines, vaccinations under the age of one and the first routine appointment may be difficult to differentiate but a minimum gap of four weeks can be applied. Children at risk can receive their second MMR dose at the age of 15 months. Hence, it has to be ensured that these early vaccinations do not get cleaned out as false recording. Figure 1.C shows the probability density of having received an MMR vaccine at a certain age and illustrates the phenomenon of some early MMR vaccinations at the age of 15 months (around 450 days). Applying only a minimum gap of four months between doses can help to include these earlier vaccinations.

**Figure 1.**
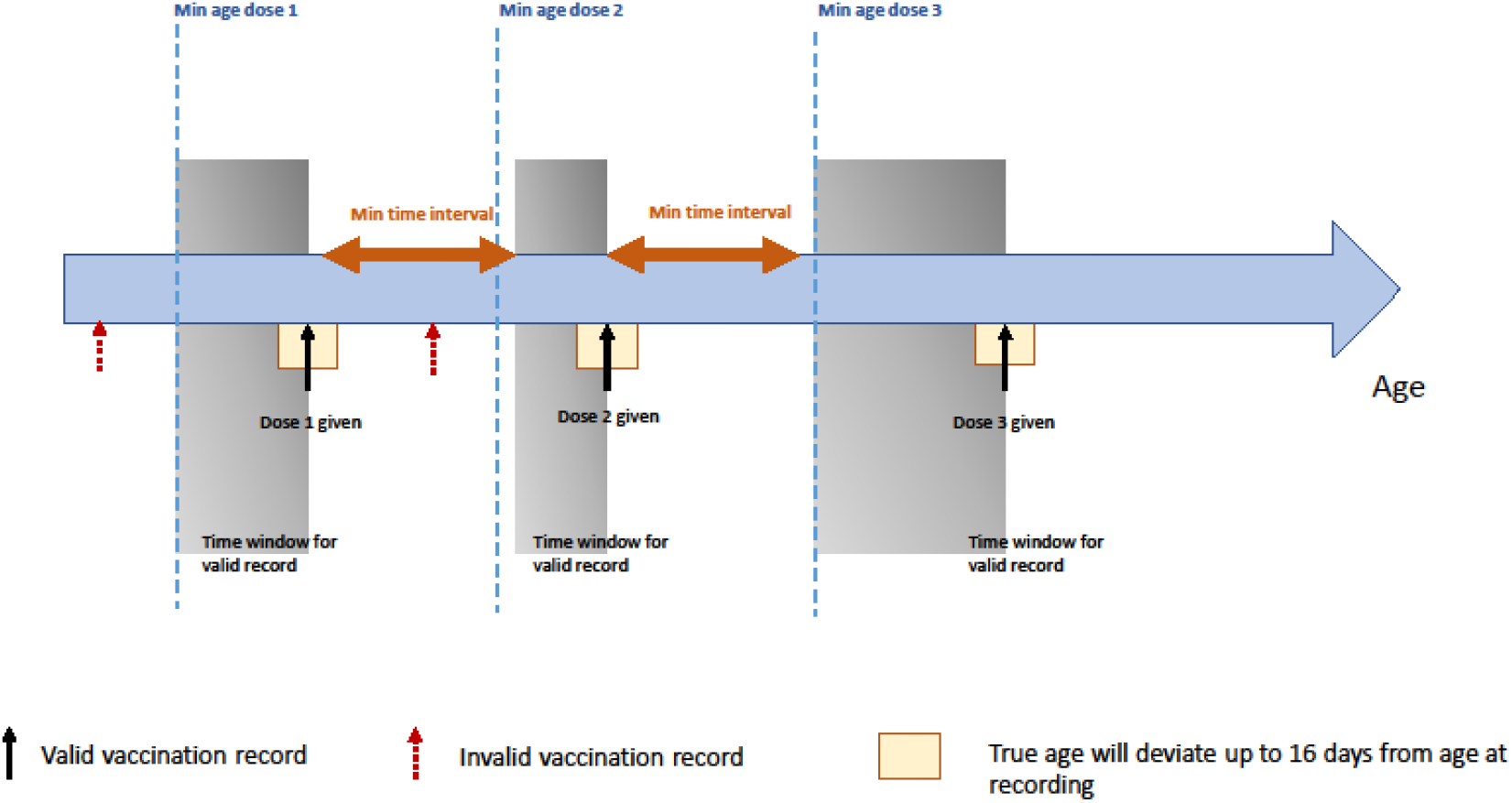
Overview of the different steps in the data cleaning algorithm after ambiguous have been already interpreted as described above. A vaccination record was interpreted as vaccine dose if given after a pre-defined minimum age and applying a minimum time interval between two doses.

**Figure 2.**
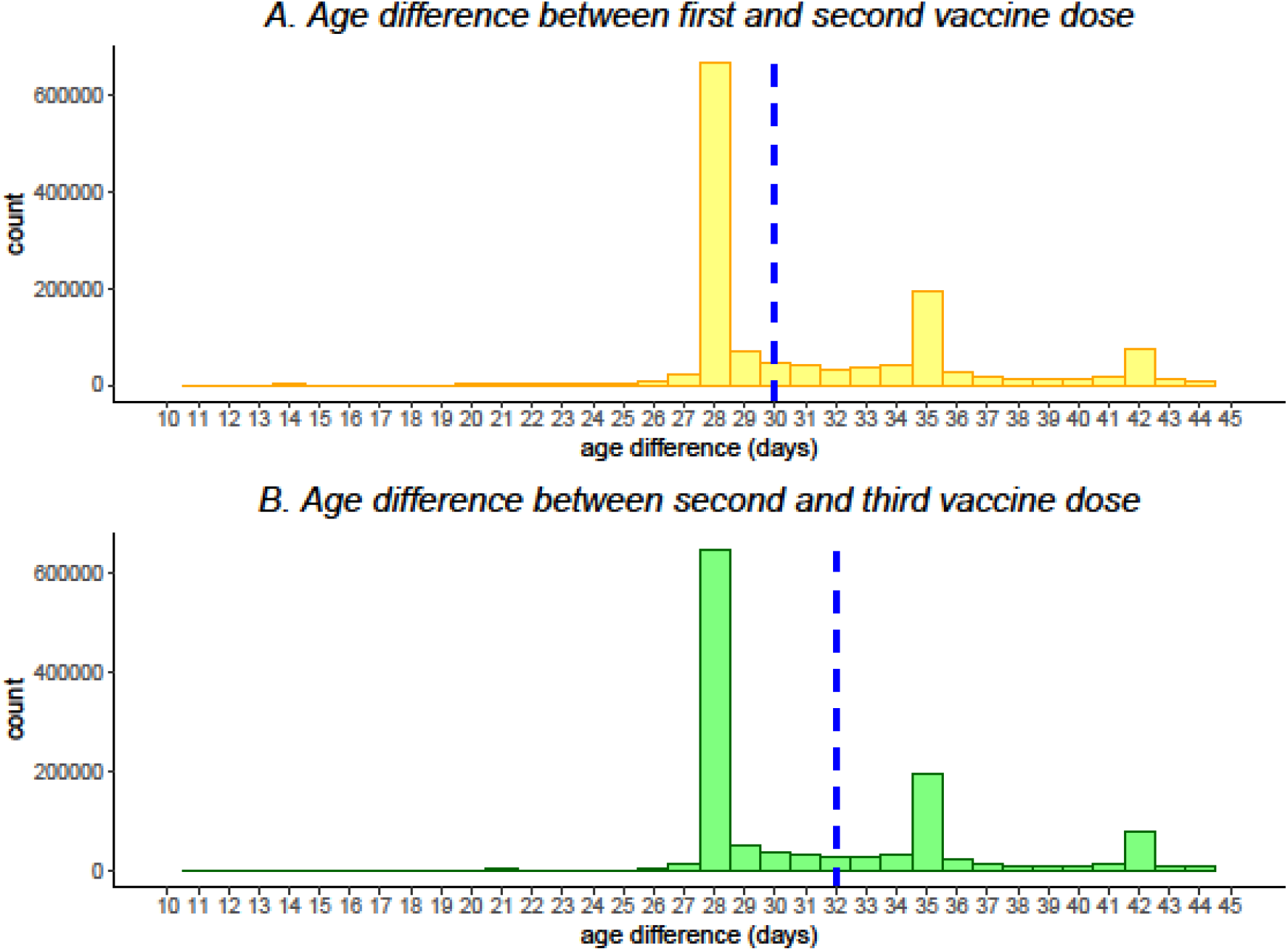
Cumulative number of vaccine events excluded from the study for different minimum age intervals between doses on the example of the DTP vaccine. The blue dashed line indicates the median age difference.

**Figure 3.**
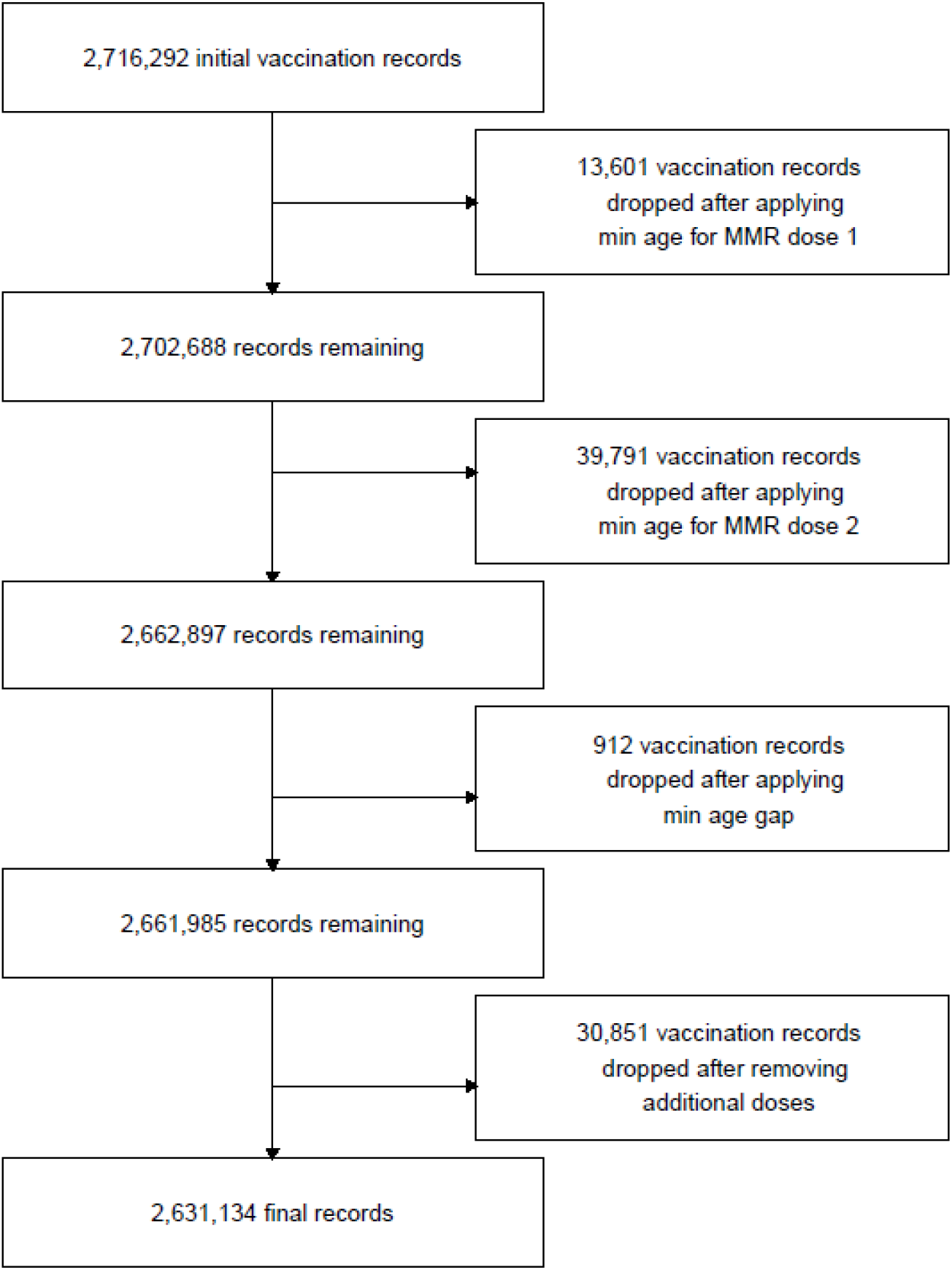
Impact of applying the vaccination algorithm to the example of the MMR vaccine. MMR: measles, mumps, rubella vaccine.

### Recommendations

- *Special indications for immunisations have to be considered with respect to study focus*
- *For studies on routine immunisations, minimum ages as demonstrated in our algorithm can be helpful to filter out additional vaccinations outside of the schedule*

## Discussion/Conclusion

Overall, in this paper we demonstrated the importance of considering the complexity of vaccines and vaccine recording in EHR when designing vaccine studies using these datasets. On the example of a cohort from CPRD Aurum in England, we showed how different methods of identifying vaccines EHR affect estimates of vaccine uptake.

In-depth knowledge of the vaccine delivery systems is required to ensure that the right data source is chosen and to consider potential biases in data collection and reporting. Vaccine schedules can change over time and special situations such as outbreaks or exposure to different pathogens might impact when a vaccine was given and have to be taken into account. This approach must be adapted to the country of the study accordingly.

When creating a code list in order to identify vaccination events in EHR, a broader approach should be taken instead of focusing on vaccine prescriptions only. We showed that using an algorithm based on a combination of prescription codes, codes indicating a vaccine refusal, neutral vaccine codes and codes for given vaccines can show similar results to national vaccine coverage estimates if wash-out periods and minimum age gaps between the vaccine codes are applied (see table S2.). Nevertheless, there is some diversity in clinical coding which has to be considered when defining these categories and might not generally apply to all primary care practices even when using the same coding software (35).

This extensive data cleaning is necessary to avoid double counting of vaccine events or lagged recording in EHR and to interpret records which are unclear as to whether the vaccine was administered. Considering more general vaccine for childhood immunisation may help to identify declined vaccinations.

We explain the slightly higher uptake of some vaccines might be due to selection bias in our study population which requires a constant registration with the same GP practice during follow-up, and hence, only captures children who did not move house within the study period. For the lower pneumococcal vaccine uptake, we hypothesise that the coding practice of this newly introduced vaccine in 2006 (36) has changed over the first years after introduction, perhaps partly due to the very common velocity coding which places commonly-used codes higher in a picking list and therewith, might hinder the use of new or rarely used codes in practices (35). Furthermore, differences to the national estimates could be due to their broader definition of acceptable codes and different deduplication methods (37).

### Strengths and limitations

Capturing vaccine status accurately is essential for high-quality studies investigating vaccine effectiveness and safety. We provided a practical guide to help researchers making decisions on study design, how to identify vaccination events, and data management to support high quality studies using EHR, supported by data from a worked example. We provided recommendations for each stage of the study design and provided step-by-step instructions for the data cleaning. In our example, we managed to replicate national estimates for vaccination coverage using our algorithm with only minor deviations.

Key limitations include the focus of this study on the UK health care system. Immunisation schedules, delivery settings and coding are likely to differ between countries. However, our algorithm framework offers an approach which can be adapted to the setting. Depending on the data set used, some information might not be accessible. In our example, there is 16-day uncertainty around the actual age at vaccination due to suppression of date of birth in CPRD. If a vaccination event was recorded more than once, it is also not possible to determine which of the events was the true vaccination event and which one might have been a consultation only. Our suggested approach using primary care records does not account for vaccines which were given outside of a primary care setting, e.g., vaccines in private clinics, BCG vaccines in hospitals, or emergency tetanus vaccinations after injury or potential exposure.

### Key recommendations for researchers

Overall, we recommend for researchers to explore the setting and the delivery pathway for vaccines first at the planning stage of the study and before deciding on a data set. After choosing an appropriate data set, researchers in a UK setting should not use prescription data only but a combination of different vaccine-related codes provided in the EHR. Antigens which are usually administered together could serve as proxy for each other if special indications for certain antigens are considered. Vaccine schedules and recommendations as well as delivery pathways undergo frequent changes which require up-to-date information for the respective study period and setting.

### Policy recommendations

We recommend for policy makers, a more centralised system for collecting vaccination data in order to bridge the gap between vaccines administered in different setting of care. Linkage with medical records would help to improve vaccine surveillance and signal detection of adverse events in children and to explore vaccine coverage in groups of special need and high-risk groups. There have been attempts to bring more vaccine data and general health data together. For children in England, the Child Health Information Services (CHIS) have been set up to collect local clinical care records of all children in an area including immunisations but do not include wider clinical data which would be necessary for safety studies or to study specific risk groups, and cannot link children who move between different regions (38). Even less data is available for people over 19 years. Other vaccine-specific data collection services such as The National Immunisation Management Service (NIMS) have initially focused on collecting data on COVID-19 and the influenza vaccine only (39).

The introduction of childhood immunizations into the Quality and Outcomes Framework (QOF) in 2021 might help to improve the data quality on conflicting or ambiguous coding (40).

Overall, a more centralised and nation-wide data collection system is needed for linking vaccination records from different sources together with relevant health care data.

## Data Availability

The study uses data from the Clinical Practice Research Datalink (CPRD). CPRD does not allow the sharing of patient-level data. The data specification for the CPRD data set is available at: https://cprd.com/cprd-aurum-may-2022-dataset. The code lists can be found at: https://github.com/Eyedeet/vaccine_methods_ehr_public/tree/main/codelists.

## Acknowledgements

This work uses data provided by patients and collected by the NHS as part of their care and support. Using patient data is vital to improve health and care for everyone. There is huge potential to make better use of information from people’s patient records, to understand more about disease, develop new treatments, monitor safety, and plan NHS services. Patient data should be kept safe and secure, to protect everyone’s privacy, and it’s important that there are safeguards to make sure that it is stored and used responsibly. Everyone should be able to find out about how patient data is used.

We would like to thank Jennie Johnson and the PRIMIS team for their insights on clinical coding practices.

## Ethics statement

We received data governance approval from CPRD (protocol number 22_001706) and ethical approval from the London School of Hygiene and Tropical Medicine’s research ethics committee (reference number 27651).

## REFERENCES

1. World Health Organization. Immunization coverage [Internet]. 2021 [cited 2022 May 9]. Available from: https://www.who.int/news-room/fact-sheets/detail/immunization-coverage

2. Galles NC, Liu PY, Updike RL, Fullman N, Nguyen J, Rolfe S, et al. Measuring routine childhood vaccination coverage in 204 countries and territories, 1980–2019: a systematic analysis for the Global Burden of Disease Study 2020, Release 1. Lancet [Internet]. 2021 Aug;398(10299):503–21. Available from: https://linkinghub.elsevier.com/retrieve/pii/S0140673621009843

3. McDonald HI, Tessier E, White JM, Woodruff M, Knowles C, Bates C, et al. Early impact of the coronavirus disease (COVID-19) pandemic and physical distancing measures on routine childhood vaccinations in England, January to April 2020. Eurosurveillance [Internet]. 2020 May 14;25(19). Available from: https://www.eurosurveillance.org/content/10.2807/1560-7917.ES.2020.25.19.2000848

4. Firman N, Marszalek M, Gutierrez A, Homer K, Williams C, Harper G, et al. Impact of the COVID-19 pandemic on timeliness and equity of measles, mumps and rubella vaccinations in North East London: a longitudinal study using electronic health records. BMJ Open [Internet]. 2022 Dec 1;12(12):e066288. Available from: https://bmjopen.bmj.com/lookup/doi/10.1136/bmjopen-2022-066288

5. de Figueiredo A, Simas C, Karafillakis E, Paterson P, Larson HJ. Mapping global trends in vaccine confidence and investigating barriers to vaccine uptake: a large-scale retrospective temporal modelling study. Lancet [Internet]. 2020 Sep 26;396(10255):898–908. Available from: https://doi.org/10.1016/S0140-6736(20)31558-0

6. Larson HJ, Jarrett C, Eckersberger E, Smith DMD, Paterson P. Understanding vaccine hesitancy around vaccines and vaccination from a global perspective: A systematic review of published literature, 2007–2012. Vaccine [Internet]. 2014 Apr;32(19):2150–9. Available from: https://linkinghub.elsevier.com/retrieve/pii/S0264410X14001443

7. Crocker-Buque T, Edelstein M, Mounier-Jack S. Interventions to reduce inequalities in vaccine uptake in children and adolescents aged <19 years: a systematic review. J Epidemiol Community Health [Internet]. 2017 Jan;71(1):87–97. Available from: https://jech.bmj.com/lookup/doi/10.1136/jech-2016-207572

8. Herrett E, Gallagher AM, Bhaskaran K, Forbes H, Mathur R, Staa T van, et al. Data Resource Profile: Clinical Practice Research Datalink (CPRD). Int J Epidemiol. 2015;44(3):827–36.

9. Wolf A, Dedman D, Campbell J, Booth H, Lunn D, Chapman J, et al. Data resource profile: Clinical Practice Research Datalink (CPRD) Aurum. Int J Epidemiol. 2019;48(6):1740–1740G.

10. Osam CS, Pierce M, Hope H, Ashcroft DM, Abel KM. The influence of maternal mental illness on vaccination uptake in children: a UK population-based cohort study. Eur J Epidemiol [Internet]. 2020;35(9):879–89. Available from: http://ovidsp.ovid.com/ovidweb.cgi?T=JS&PAGE=reference&D=medl&NEWS=N&AN=32328992

11. Peppa M, Thomas SL, Minassian C, Walker JL, McDonald HI, Andrews NJ, et al. Seasonal Influenza Vaccination During Pregnancy and the Risk of Major Congenital Malformations in Live-born Infants: A 2010–2016 Historical Cohort Study. Clin Infect Dis. 2020;(Xx Xxxx):1–9.

12. Pottegård A, Lund LC, Karlstad Ø, Dahl J, Andersen M, Hallas J, et al. Arterial events, venous thromboembolism, thrombocytopenia, and bleeding after vaccination with Oxford-AstraZeneca ChAdOx1-S in Denmark and Norway: population based cohort study. BMJ [Internet]. 2021 May 5;n1114. Available from: https://www.bmj.com/lookup/doi/10.1136/bmj.n1114

13. Walker JL, Rentsch CT, McDonald HI, Bak J, Minassian C, Amirthalingam G, et al. Social determinants of pertussis and influenza vaccine uptake in pregnancy: a national cohort study in England using electronic health records. BMJ Open. 2021;11(6):e046545.

14. Thomas SL, Walker JL, Fenty J, Atkins KE, Elliot AJ, Hughes HE, et al. Impact of the national rotavirus vaccination programme on acute gastroenteritis in England and associated costs averted. Vaccine [Internet]. 2017 Jan;35(4):680–6. Available from: https://linkinghub.elsevier.com/retrieve/pii/S0264410X16311197

15. Andrews N, Stowe J, Kuyumdzhieva G, Sile B, Yonova I, de Lusignan S, et al. Impact of the herpes zoster vaccination programme on hospitalised and general practice consulted herpes zoster in the 5 years after its introduction in England: a population-based study. BMJ Open [Internet]. 2020 Jul 7;10(7):e037458. Available from: https://bmjopen.bmj.com/lookup/doi/10.1136/bmjopen-2020-037458

16. Smeeth L, Cook C, Fombonne E, Heavey L, Rodrigues LC, Smith PG, et al. MMR vaccination and pervasive developmental disorders: a case-control study. Lancet (London, England) [Internet]. 2004;364(9438):963–9. Available from: http://ovidsp.ovid.com/ovidweb.cgi?T=JS&PAGE=reference&D=med5&NEWS=N&AN=15364187

17. Walker JL, Andrews NJ, Atchison CJ, Collins S, Allen DJ, Ramsay ME, et al. Effectiveness of oral rotavirus vaccination in England against rotavirus-confirmed and all-cause acute gastroenteritis. Vaccine X [Internet]. 2019 Apr;1:100005. Available from: https://linkinghub.elsevier.com/retrieve/pii/S2590136219300026

18. Shortreed SM, Cook AJ, Coley RY, Bobb JF, Nelson JC. Challenges and Opportunities for Using Big Health Care Data to Advance Medical Science and Public Health. Am J Epidemiol [Internet]. 2019 May 1;188(5):851–61. Available from: https://academic.oup.com/aje/article/188/5/851/5381891

19. Clinical Practice Research Datalink. CPRD Aurum May 2022 data set [Internet]. CPRD Aurum May 2022 (Version 2022.05.001) [data set]. 2022 [cited 2023 Feb 13]. Available from: https://cprd.com/cprd-aurum-may-2022-dataset

20. Herrett E, Thomas SL, Schoonen WM, Smeeth L, Hall AJ. Validation and validity of diagnoses in the General Practice Research Database: a systematic review. Br J Clin Pharmacol [Internet]. 2010 Jan;69(1):4–14. Available from: https://onlinelibrary.wiley.com/doi/10.1111/j.1365-2125.2009.03537.x

21. Medicines & Healthcare products Regulatory Agency. Clinical practice research data link - linked data [Internet]. 2021. Available from: https://www.cprd.com/linked-data

22. Llamas A, Amirthalingam G, Andrews N, Edelstein M. Delivering prenatal pertussis vaccine through maternity services in England: What is the impact on vaccine coverage? Vaccine [Internet]. 2020 Jul;38(33):5332–6. Available from: https://linkinghub.elsevier.com/retrieve/pii/S0264410X20307234

23. Tiley K, Tessier E, White JM, Andrews N, Saliba V, Ramsay M, et al. School-based vaccination programmes: An evaluation of school immunisation delivery models in England in 2015/16. Vaccine. 2020;38(15):3149–56.

24. UK Health Security Agency. Flu vaccination programme 2022 to 2023: briefing for primary schools. [Internet]. 2022 [cited 2023 Feb 26]. Available from: https://www.gov.uk/government/publications/flu-vaccination-in-schools/flu-vaccination-programme-2021-to-2022-briefing-for-schools#the-nasal-flu-vaccine

25. UK Health Security Agency. 19 Influenza. In: Immunisation against infectious disease (Green Book) [Internet]. 2020. Available from: https://www.gov.uk/government/publications/influenza-the-green-book-chapter-19

26. Crocker-Buque T, Edelstein M, Mounier-Jack S. A process evaluation of how the routine vaccination programme is implemented at GP practices in England. Implement Sci [Internet]. 2018 Dec 22;13(1):132. Available from: https://implementationscience.biomedcentral.com/articles/10.1186/s13012-018-0824-8

27. Medicines & Healthcare products Regulatory Agency. Patient group directions: who can use them [Internet]. 2017 [cited 2023 Feb 13]. Available from: https://www.gov.uk/government/publications/patient-group-directions-pgds/patient-group-directions-who-can-use-them

28. UK Health Security Agency. 11 The UK immunisation schedule. In: Immunisation against infectious disease (Green Book) [Internet]. London; 2020. Available from: https://www.gov.uk/government/collections/immunisation-against-infectious-disease-the-green-book

29. UK Health Security Agency. 21 Measles. In: The Green Book [Internet]. 2019. Available from: https://www.gov.uk/government/publications/measles-the-green-book-chapter-21

30. Pesco P, Bergero P, Fabricius G, Hozbor D. Mathematical modeling of delayed pertussis vaccination in infants. Vaccine [Internet]. 2015;33(41):5475–80. Available from: http://dx.doi.org/10.1016/j.vaccine.2015.07.005

31. Stowe J, Andrews N, Ladhani S, Miller E. The risk of intussusception following monovalent rotavirus vaccination in England: A self-controlled case-series evaluation. Vaccine [Internet]. 2016 Jul;34(32):3684–9. Available from: https://linkinghub.elsevier.com/retrieve/pii/S0264410X16302043

32. UK Health Security Agency. 24 Pertussis [Internet]. The Green Book. 2016. Available from: UK Health Security Agency

33. NHS Digital & UK Health Security Agency. Childhood Vaccination Coverage Statistics - 2020-21 [Internet]. 2021 [cited 2021 Oct 12]. Available from: https://digital.nhs.uk/data-and-information/publications/statistical/nhs-immunisation-statistics/england---2020-21

34. UK Health Security Agency. 30 Tetanus [Internet]. The Green Book. 2022 [cited 2022 Nov 23]. Available from: https://www.gov.uk/government/publications/tetanus-the-green-book-chapter-30

35. Waize T, Anandarajah S, Dhoul N, De Lusignan S. Variation in clinical coding lists in UK general practice: a barrier to consistent data entry? J Innov Heal Informatics. 2007 Sep;15(3):143–50.

36. UK Health Security Agency. 25 Pneumococcal [Internet]. Immunisation against infectious disease (Green Book). 2020 [cited 2022 Jun 14]. Available from: https://www.gov.uk/government/publications/pneumococcal-the-green-book-chapter-25

37. PRIMIS team. National vaccinatio uptake programmes [Internet]. [cited 2023 Feb 28]. Available from: https://www.nottingham.ac.uk/primis/projects/national-vaccination-programmes.aspx

38. Local Government Assocication. Child Health Informaiton Services [Internet]. 2022 [cited 2022 Nov 14]. Available from: https://www.local.gov.uk/topics/social-care-health-and-integration/public-health/children-public-health-transfer/child-health-information-services

39. NHS England. National Vaccination programmes [Internet]. 2022 [cited 2022 Dec 21]. Available from: https://www.england.nhs.uk/contact-us/privacy-notice/national-flu-vaccination-programme/

40. NHS Digital. Quality and Outcomes Framework, 2021-22 [Internet]. 2022 [cited 2022 Nov 16]. Available from: https://digital.nhs.uk/data-and-information/publications/statistical/quality-and-outcomes-framework-achievement-prevalence-and-exceptions-data/2021-22

