## Supplementary material for "Methodological challenges and recommendations for identifying childhood immunisations using routine electronic health records in the United Kingdom"

### Appendix

*Table S1. Cohort demographics for children born between January 2006 and December 2014 in England.*

|  | **Cohort without minimum follow up (N = 1,735,692)** | | **Cohort with follow up to age 1 (N =775,567 )** | | **Cohort with follow up to age 2 ( N = 704,481)** | | **Cohort with follow up to age 5 (N = 573,015)** | |
| --- | --- | --- | --- | --- | --- | --- | --- | --- |
|  | N | % | N | % | N | % | N | % |
| **Sex** | | | | | | | | |
| male | 890150 | 51.29 | 398854 | 51.35 | 362130 | 51.33 | 294501 | 51.34 |
| female | 845521 | 48.71 | 377851 | 48.65 | 343295 | 48.66 | 279177 | 48.66 |
| intersex | 21 | <0.01 | 3 | <0.01 | 3 | <0.01 | 3 | <0.01 |
| **Ethnicity** | | | | | | | | |
| White | 978162 | 56.36 | 462606 | 59.56 | 422458 | 59.89 | 347705 | 60.61 |
| Asian/British Asian | 142181 | 8.19 | 59332 | 7.64 | 54313 | 7.7 | 43894 | 7.65 |
| Black/Black British | 95773 | 5.52 | 32886 | 4.23 | 28603 | 4.05 | 20888 | 3.64 |
| Mixed | 37517 | 2.16 | 12443 | 1.6 | 10892 | 1.54 | 8113 | 1.41 |
| Other | 57386 | 3.31 | 21690 | 2.79 | 18982 | 2.69 | 14275 | 2.49 |
| Unknown/missing | 424673 | 24.47 | 187751 | 24.17 | 170180 | 24.12 | 138806 | 24.20 |
| **Year of birth** | | | | | | | | |
| 2006 | 16572 | 0.95 | 6821 | 0.88 | 6257 | 0.89 | 5262 | 0.92 |
| 2007 | 222822 | 12.84 | 88426 | 11.38 | 80902 | 11.47 | 66420 | 11.58 |
| 2008 | 232061 | 13.37 | 93589 | 12.05 | 85146 | 12.07 | 69944 | 12.19 |
| 2009 | 233629 | 13.46 | 87917 | 11.32 | 79753 | 11.31 | 64751 | 11.29 |
| 2010 | 238321 | 13.73 | 95114 | 12.25 | 86245 | 12.23 | 69793 | 12.17 |
| 2011 | 226220 | 13.03 | 98652 | 12.7 | 89322 | 12.66 | 72508 | 12.64 |
| 2012 | 213537 | 12.3 | 102732 | 13.23 | 92765 | 13.15 | 75451 | 13.15 |
| 2013 | 186655 | 10.75 | 100566 | 12.95 | 91338 | 12.95 | 74626 | 13.01 |
| 2014 | 165875 | 9.56 | 102891 | 13.25 | 93700 | 13.28 | 74926 | 13.06 |
| **Region** | | | | | | | | |
| North East | 47309 | 2.73 | 24108 | 3.1 | 22395 | 3.17 | 19187 | 3.34 |
| North West | 286418 | 16.5 | 141236 | 18.18 | 131191 | 18.6 | 112412 | 19.59 |
| Yorkshire & The Humber | 57249 | 3.3 | 27271 | 3.51 | 25125 | 3.56 | 21157 | 3.69 |
| East Midland | 36193 | 2.09 | 16671 | 2.15 | 15228 | 2.16 | 12687 | 2.21 |
| West Midlands | 265142 | 15.28 | 121950 | 15.7 | 111838 | 15.85 | 93450 | 16.29 |
| East of England | 77026 | 4.44 | 35197 | 4.53 | 32243 | 4.57 | 26728 | 4.66 |
| London | 431414 | 24.86 | 167600 | 21.58 | 145650 | 20.65 | 104892 | 18.28 |
| South East | 347887 | 20.04 | 157143 | 20.23 | 143441 | 20.33 | 118131 | 20.59 |
| South West | 187054 | 10.78 | 85532 | 11.01 | 78317 | 11.1 | 65037 | 11.34 |

##### Table S.2 Number of vaccination-related codes identified by different category using antigen specific code lists versus more general code lists.

|  | **Vaccine administered** | **Neutral vaccine code** | **Vaccine product code** | **Vaccine declined** |
| --- | --- | --- | --- | --- |
| DTP specific code lists | 3,200,972 | 5,677,041 | 57,804 | 12,682 |
| General vaccine code lists only | 55,138 | 35,761 | - | 35,761 |
| Combination of DTP specific terms and general terms | 3,256,110 | 6,837,603 | 57,804 | 48,443 |

**Note: The code lists for the DTP vaccine and the different categories are shared with this paper as an additional supplement.**

##### Table S.3 Number of overall received vaccine doses identified by using either antigen-specific code list or a code list including more general vaccination terms

|  | Only DTP specific codes  (N= 1,667,450) | DTP and general codes  (total = 1,657,868) |
| --- | --- | --- |
| One DTP dose | 40,639 (2.44%) | 40,921 (2.47%) |
| Two DTP doses | 39,584 (2.37%) | 38,628 (2.33%) |
| Three DTP doses | 509,526 (30.56%) | 477,900 (28.82%) |
| Four DTP doses | 1,070,032 (64.17%) | 1,073,609 (64.76%) |
| All vaccines declined | 7,669 (0.4%) | 26,810 (1.62%) |

##### Table S.4 Potential combination of vaccine related records for an individual on the same day, the interpretation of their combination, and actual number of record combinations occurring on the example of MMR vaccine.

|  | **Vaccine administered** | **Neutral vaccine code** | **Vaccine product code** | **Vaccine declined** |
| --- | --- | --- | --- | --- |
| **Vaccine administered** | Vaccinated  1,358,852 (49.90%) | Vaccinated  26,166 (0.96%) | Vaccinated  14,108 (0.52%) | Conflict  327 (0.01%) |
| **Neutral vaccine code** | - | Vaccinated  1,290,947 (47.41%) | Vaccinated  11,912 (0.44%) | Unvaccinated  448 (0.02%) |
| **Vaccine product code** | - | - | Vaccinated  331( 0.01%) | Conflict  0 (0.00%) |
| **Vaccine declined** | - | - |  | Unvaccinated  20,024 (0.74%) |

**Note: The denominator for the proportions is the overall sum of days with any type of vaccination records.**

##### Table S5 Summarising the overall number of events classified as vaccinated, vaccine declined or conflicting evidence

|  | Number of events of vaccination (%) | Number of events of vaccination declined (%) | Number of conflicting events (%) |
| --- | --- | --- | --- |
| DTP vaccine | 6101360 (99.84%) | 9831 (0.16%) | 138 (<0.00%) |
| MMR vaccine | 2,702,306 (99.24%) | 20,472 (0.75%) | 337 (<0.00%) |
| Pneumococcal vaccine | 4,586,379 (99.87%) | 5,815 (0.13%) | <5 (<0.00%) |

##### S.6: Description of the algorithm to identify vaccination events

To differentiate between different vaccination events and avoid double counting of the same events, we propose the following stages before determining the number of vaccine doses:

1. We established how many children had a vaccination recorded around their potential date of birth (16 days before and after our estimated date). These children were excluded from the analysis as it had to be assumed that their date of birth or their vaccination timing were not recorded correctly.
2. For each vaccine dose, we graphically described the uptake by date to determine a sensible minimum age threshold for each dose. This minimum age was also compared to and informed by the recommendations of the Green Book (UK Health Security Agency, 2020a).
3. For two vaccine doses of the same antigen, we calculated the age difference between the two doses to determinate a sensible minimum time interval between two doses. The minimum time interval was also compared to and informed by the recommended minimum interval in the national guidance (the ‘Green Book’).
4. We then applied this minimum age thresholds and time intervals between doses.

Each of these steps will be now illustrated on the example of a pertussis-antigen containing vaccine.

Initially, 1,696,896 children in the study record had any health record related to the pertussis vaccination.

Stage 1 – checking date of birth relative to vaccination record

After defining vaccination events, the date of birth in relation to a first vaccination record was checked. 3,240 children (0.01% of all children with a record) had their first pertussis vaccine recorded within two weeks of their potential date of birth. Of these events with conflicting timing, 77.56% were recorded within the same year of the observation date which makes backdating as a potential error course more unlikely. We also inspected the year of birth of these children to examine whether some years came along with poorer recording. As there was no obvious pattern by calendar year, we excluded all these children from the study population, assuming that the quality of their data recording was not sufficient.

Step 2 – describing the uptake of each dose by age and defining a minimum age

The age of uptake for each vaccine dose was graphically and numerically described (see table 6 and figure 1). The minimum age for each dose was chosen considering the recommended minimum age in the Green Book (UK Health Security Agency, 2016), the observed rise in the count of recorded uptake by age (see figure 1) and the youngest possible age following the vaccination schedule and allowing 16 days of uncertainty regarding the date of birth.

In our example, the observed rise in the recording of vaccine uptake over age aligned well with the expected minimum age for each dose. We decided to allow for two more days of imprecise recording to avoid under-recording of doses which might have been given minimally too early because of GP opening times or other practical reasons. This resulted in minimum age of 38, 66, and 96 days respectively for the first three pertussis doses. The minimum age of the fourth dose was set at 1077 days which aligned with the youngest possible age for a pertussis booster as recommended in the Green Book (UK Health Security Agency, 2016).

##### Table S.7 Age in days for the uptake of different pertussis vaccine doses and time in days to the next vaccination record.

|  | Median | IQR | Min | Max | Expected age range |
| --- | --- | --- | --- | --- | --- |
| First pertussis dose | 64 | 55-73 | 16 | 1827 | 40-72 |
| Second pertussis dose | 97 | 87-111 | 18 | 1827 | 68-100 |
| Time to next vaccination records (dose 1 and 2) | 30 | 28-39 | 1 | 1777 | 28 |
| Third pertussis dose | 131 | 119-152 | 41 | 1827 | 98-128 |
| Time to next vaccination record (dose 2 and 3) | 32 | 28-42 | 1 | 1745 | 28 |
| Fourth pertussis dose | 1273 | 1239-1328 | 60 | 1827 | 1199-1231 |
| Time to next vaccination record (dose 3 and 4) | 1135 | 1105-1188 | 1 | 1739 | - |

5,770 children (0.34% of all children with a pertussis vaccination record) did not fulfil the minimum age for the first pertussis dose, 3,522 (0.21%) for the second dose, 3,219 (0.19%) for the third dose and 30,874 (1.82%) for the preschool booster.

Step 3 – Defining a minimum age gap between doses

After looking at the uptake by age for each individual, we calculated the age difference between two consequential doses. The Green Books recommends a minimum of at least 4 weeks between two doses (UK Health Security Agency, 2016) which was well reflected in the data showing a median age difference of 28 days between dose one and two, and dose two and three for study population (see table S.5). For consistency, we also allowed an imprecision of two days for the recording between two consequential doses which resulted in a minimum age gap of 26 days between dose one and two, and dose two and three.

For the gap between dose three and four, a different minimum gap was chosen because the Green Book recommend at least one year break between completion of the primary course and the preschool booster (UK Health Security Agency, 2016). Accordingly, we set the minimum age gap between third and fourth dose of pertussis vaccine to 363 days (including two days of imprecision of recording).

1.14% of the children with record did not have the minimum age gap between pertussis vaccine dose one and two, 1.01% did not have the minimum gap between dose two and three, and 1.59% did not have the minimum gap between dose three and the preschool booster.

Step 4 – applying minimum ages and minimum age gaps to the different vaccine doses

After the defining a minimum age for each dose and a minimum age gap between the different doses, the following algorithm can be applied.

6. Order every individual’s observations chronologically, count the overall number of vaccine events per individual and number them consecutively.
7. Discard all vaccine events for the first pertussis dose which do not comply with the minimum age of 38 days. Repeat step one. Repeat step two until no more events get dropped.
8. Discard all vaccine events for the second pertussis dose which do not comply with the minimum age of 66 days. Repeat step one. Repeat step three until no more events get dropped.
9. Discard all vaccine events for the second pertussis dose if they do not comply with the minimum age gap of 26 days. Repeat step one. Repeat step four until no more events get dropped.
10. Discard all vaccine events for the third pertussis dose which do not comply with the minimum age of 96 days. Repeat step one. Repeat step five until no more events get dropped.
11. Discard all vaccine events for the third pertussis dose if they do not comply with the minimum age gap of 26 days. Repeat step one. Repeat step six until no more events get dropped.
12. Discard all vaccine events for the fourth pertussis dose which do not comply with the minimum age of 1077 days. Repeat step one. Repeat step seven until no more events get dropped.
13. Discard all vaccine events for the fourth pertussis dose if they do not comply with the minimum age gap of 363 days. Repeat step one. Repeat step eight until no more events get dropped.

##### Table S.8 Recommendations for minimum age and age gaps for different vaccines

| Item | DTP vaccine | MMR vaccine | PCV vaccine |
| --- | --- | --- | --- |
| Min. age for dose 1 | 38 days | 437 days | 38 days |
| Age gap between dose 1 and 2 | 26 days | 26 days | 26 days |
| Min. age for dose 2 | 66 days | 530 days | 96 days |
| Age gap between dose 2 and 3 | 26 days | NA | 26 days |
| Min. age for dose 3 | 96 days | NA | 347 days |
| Age gap between dose 3 and 4 | 363 days | NA | NA |
| Min. Age for dose 4 | 1077 days | NA | NA |

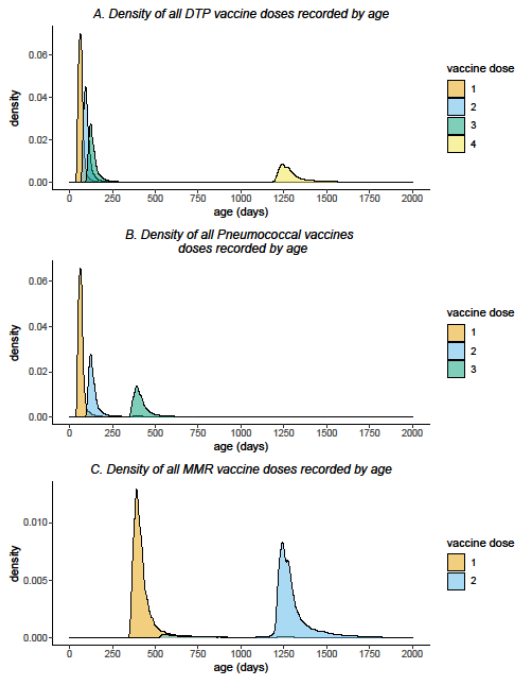

**Figure S9. Probability density of recording a vaccine dose by age for the DTP vaccine (A), the pneumococcal vaccine (B) and the MMR vaccine (C). DTP: diphtheria, tetanus, pertussis vaccine. MMR: measles, mumps, rubella vaccine.**
